# Is Netrin-1 Associated with the Development of Fibrosis in Systemic Sclerosis?

**DOI:** 10.1101/2022.02.05.22270510

**Authors:** Yüksel Maraş, Ahmet Kor, Esra Fırat Oğuz, Alper Sarı, Kevser Gök, Ali Akdoğan

**Author notes:** **Corresponding author:** DR. Ahmet Kor, Department of Rheumatology, Ankara Yıldırım Beyazıt University Faculty of Medicine Ankara City, Universities District 1604. Street No: 9 Çankaya/ANKARA/TURKEY **Email:**.

## Abstract

**Background:** Netrin-1 is a laminin class protein that guides the axonal during the first embryonic development, has pushing and pulling properties, and has axonal chemoattractant activity. Netrin-1 has been shown to increase the effect of fibrosis in mouse lung and human SSc lung cell cultures. In this study, we aimed to investigate the relationship between Netrin-1 and Systemic sclerosis (SSc) and to emphasize the role of Netrin-1 in the pathophysiology of SSc by increasing the known VEGF and M2 macrophage expression, which supports the fibrotic process.

**Methods:** The study included 56 SSc patients with a mean age of 48·.08±13.59 years and 58 healthy volunteers with a mean age of 48.01±11.59 years. SSc organ involvements were scanned retrospectively from patient files, and patients were grouped according to SSc complications. Calculation of Netrin-1 levels was performed using a quantitative sandwich enzyme immunoassay method with an ELISA kit (Elabscience, Texas, USA; catalog number: E-EL-H2328; lot number: GZWTKZ5SWK). The modified Rodnan skin score (mRSS) was used for skin thickness scoring in SSc patients.

**Results:** The median value of Netrin-1 was found to be significantly higher in SSc (268.8 [82.75-1006.64]) than in controls (108.63 [21.02-351.49]) (p<0.0001). In ROC analysis, a cut-off value of 354.24 for Netrin in SSc was found to provide a sensitive confidence interval with 32.8% sensitivity and 98.3% specificity (AUC[95% CI]: 0.746-0.895, p<0.0001). There was no significant correlation between Netrin-1 level, organ involvement in SSc, and mRSS (p>0.05).

**Conclusion:** We found a significant relationship between Netrin-1 levels and SSc disease in this study. Our study is the first clinical study in which Netrin-1 elevation was demonstrated in SSc patients.

## Introduction

Systemic sclerosis (SSc), often called scleroderma, is an autoimmune, destructive systemic connective tissue disease characterized by organ fibrosis and vasculopathy. Pathophysiological mechanisms that may play a role in disease development include platelet activation, fibroblast proliferation, endothelial disruption, fetal microchimerism, and increased transforming growth factor-β. In addition, VEGF is an essential signaling factor contributing to the pathogenesis of SSc, even in the earliest clinically detectable stages of the disease [1].

Netrin-1 is a laminin class protein that provides axonal guidance during the first embryonic development and has axonal chemoattractant activity by binding to UNC5 (uncoordinated-5) and DCC (Deleted in Colorectal Cancer) receptors [2]. In an animal study conducted by Aranzazu M. and et al. it was determined that when 8-week-old arthritis mice were treated with anti-Netrin-1 or anti-Unc5b antibodies for 4 weeks, there was a significant decrease in inflammation (p<0.001) and joint erosion, did not develop when compared to mice that did not receive treatment. This study also showed a reduction in cathepsin K+ and CD68+ cells and a marked reduction in osteoclasts in animals treated with anti-Netrin-1/anti-Unc5b. It has been shown that blockade of Netrin-1/Unc5b with anti-Netrin-1/anti-Unc5b antibodies prevents bone loss and reduces the severity of arthritis provoked by K/BxN serum transfer [3]. Similarly, another animal study showed that Netrin-1 is an autocrine and paracrine factor that increases osteoclast differentiation and function produced by osteoclast precursors and inflammatory cells [4]. Studies in the field of obesity have shown that the exit of macrophages from the inflammatory environment is prevented by Netrin-1, and thus Netrin-1 contributes to the development of atherosclerosis [5-7]. Unlike these results, there are also studies showing that Netrin-1 has proangiogenic [8], anti-apoptotic [9], and anti-inflammatory properties [10], as well as cardioprotective effects against myocardial damage [11], and reducing endothelial dysfunction in diabetes [12]. It is thought that Netrin-1 has proangiogenic properties by increasing VEGF expression and receptor response [11-13].

The relationship between SSc, characterized by skin and organ fibrosis, and Netrin-1 is unknown. In a limited number of studies conducted in this area, it has been shown that Netrin-1 increases the development of fibrosis in bleomycin-induced mouse lung and SSc lung cell culture [16,17]. However, no study eval plasma Netrin-1 levels in SSc and complications related to SSc. In this study, we aimed to evaluate the levels of Netrin-1 between SSc and healthy controls and to emphasize the role of the known effects of Netrin-1 in the pathophysiology of SSc, which increases VEGF and supports the fibrotic process.

## Methods

### Patients

A total of 56 SSc patients (mean age: 48.08±13.59), 53 females and 3 males, who were followed up in Ankara city hospital and Hacettepe medical school rheumatology department, diagnosed according to 2013 ACR (American College of Rheumatology)\EULAR (European League Against Rheumatism) Systemic Sclerosis classification criteria were included in the study. A total of 58 volunteers (mean age: 48.01±11.59 years) consisting of 54 females and 4 males without rheumatologic disease were selected for the control group. Organ involvements due to SSc were reviewed retrospectively from patient files. Patients with malignant disease, active infection, peripheral arterial disease, and vascular thrombosis were excluded from the study. Individuals were included in the study groups between 1.7.2020 and 1.7.2021. Modified Rodnan skin score (mRSS) was used for skin thickness scoring in SSc patients. For mRSS, 17 body areas were evaluated and scored 0-51 points in the range. All patients included in the study gave informed consent.

### Obtaining Sample Samples and Calculating Netrin-1 Values

Venous blood samples were centrifuged at 1300 x g for 10 minutes in vacuum tubes of approximately 10 mL. Samples divided into Eppendorf tubes were kept at -80 °C until analysis. Calculation of Netrin-1 levels was performed using a quantitative sandwich enzyme immunoassay method with an ELISA kit (Elabscience, Texas, USA; catalog number: E-EL-H2328; lot number: GZWTKZ5SWK).

Netrin-1 standards and serum samples added to micro ELISA plate wells and their specific antibodies were incubated for 1.5 hours at 37°C, followed by a Netrin-1 specific biotin-enriched detection antibody and set for an additional 1 hour at 37°.

A biotin-enriched detection antibody specific to Avidin-Horseradish Peroxidase (HRP) and human Netrin-1 was added and incubated at 37°C for 30 minutes. After washing the free components, substrate solution was added to each well. After this treatment, only wells containing biotin-enriched detection antibodies, human Netrin-1, and Avidin-HRP conjugate were detected in blu. The yellow color was obtained by the enzyme-substrate reaction’s termination after adding the stop solution. A microplate reader at 450 nm wavelength was used for optical density measurement by spectrophotometric measurement—optical density level increases in direct proportion to the concentration of human Netrin-1. The human Netrin-1 level in the study samples was calculated based on a comparison of the optical density standard curves of the models. The test was sensitive to detect the Netrin-1 level range of 31.25–2000 pg/mL. Inter-assay and intra-assay precision of <10% was available for all low, medium, and high Netrin-1 concentrations.

### Statistical analysis

Kolmogorov Smirnov test was used to determine the normal distribution in continuous variables. Mann Whitney UU test and Independent Samples T-Test was used to determine the statistically significant difference between the groups. Pearson correlation analysis was used to determine the correlation between study parameters. Descriptive statistics were given using mean and standard deviation (mean±SD) for normally distributed variables and median and IQR (Inter-quartile range) values for non-normally distributed variables. Comparisons between multiple groups after Bonferroni correction One-way Anova post hoc Tukey test for parametric numerical variables; Independent Samples Kruskal Wallis test was applied for nonparametric numerical variables. A Chi-square test and a Fisher test were used to compare categorical data. While testing the diagnostic accuracy measures of the indexes, ROC analysis was used, and AUC was presented with 95% confidence intervals. Youden’s index was used while determining the optimum cut-off value, and diagnostic accuracy criteria for the cut-off value were presented. The p<0·05 level was taken as the lower limit considered statistically significant. The statistical analyses were calculated using the ‘Statistical Packages for the Social sciences(SPSS) version 22.0 package program.

## Results

56 SSc patients (53 female, three male) with a mean age of 48.08±13.59 years and 58 healthy volunteers (54 female, four male) with a mean age of 48.01±11.59 were included in the study. The comparison of the groups in terms of age and gender was found to be similar (p>0.05). In the comparison between the SSc and control groups, smoking and accompanying comorbid diseases of the individuals were identical.

The comparison of the SSc and control groups in terms of study parameters are shown in Table 1. There was no significant difference between the SSc and control groups regarding hemoglobin, thrombocyte, neutrophil, creatinine, and alanine aminotransferase averages (p>0.05). The median value of C-reactive protein (CRP) was found to be significantly higher in the SSc group (0.524 [0.01-1.45]) compared to the control group (0.389 [0.2-0.794]) (p=0.032). The median value of erythrocyte sedimentation rate (ESR) was found to be significantly higher in the SSc group (14 [1-36]) compared to the control group (8 [1-28]) (p=0.043). The median value of Netrin-1 was found to be significantly higher in SSc (268.8 [82.75-1006.64]) than in controls (108.63 [21.02-351.49]) (p<0.0001). Figure 1 shows the distribution of the median values of Netrin-1 between the patient and control groups.

**Table 1.** Comparison of Study Parameters SSc and Control Groups.

| Parameter | SSc | Control | P-Value |
| --- | --- | --- | --- |
| Hemoglobin, mean ± SD [$\times 10^9/L$] | 13.11±1.19 | 13.31±1.36 | 0.23 |
| Trombosit, mean ± SD [$\times 10^9/L$] | 280.60±84.07 | 272.71±64.20 | 0.17 |
| WBC, mean ± SD [$\times 10^9/L$] | 7.43±1.88 | 6.89±1.79 | 0.11 |
| Nötrofil, mean ± SD [$\times 10^9/L$] | 5.84±2.87 | 4.39±1.81 | 0.09 |
| Creatinine, mean ± SD [mg/dl] | 0.66±0.14 | 0.70±0.11 | 0.14 |
| ALT, mean ± SD [mg/dl] | 19.86±16.61 | 23.87±9.76 | 0.19 |
| CRP, (IQR) [mg/dl] | 0.524 (0.01-1.45) | 0.389 (0.2-0.79) | 0.032 |
| ESR, (IQR) [mm/h] | 14 (1-36) | 8 (1-28) | 0.043 |
| Netrin-1, (IQR) [pg/mL] | 268.8 (82.75-1006.64) | 108.63 (21.02-351.49) | <0.0001 |

CRP: C-reactive protein, ESR: Erythrocyte Sedimentation Rate, WBC: White blood cells, ALT: Alanine aminotransferase

**Fig. 1.**
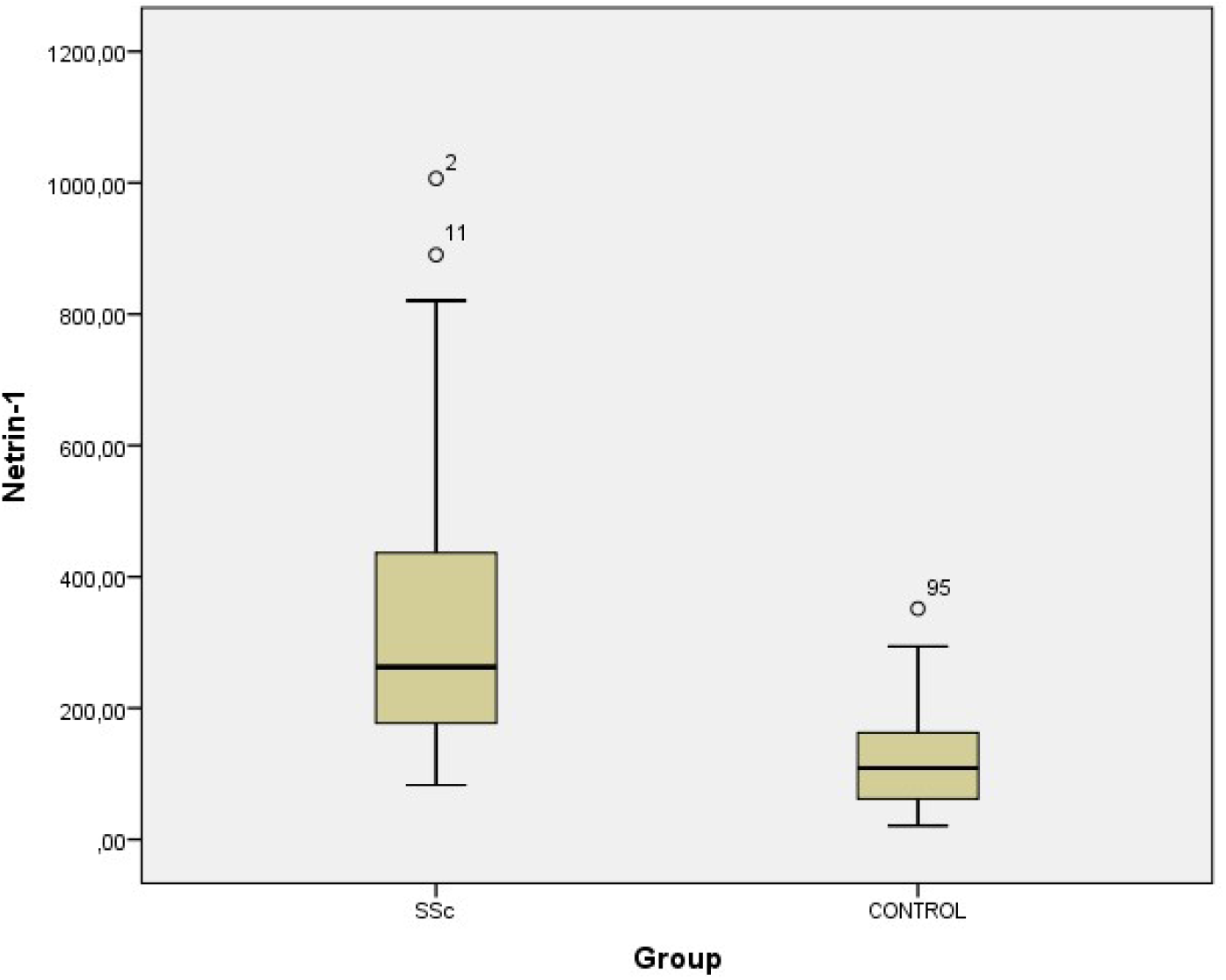
Display of Netrin-1 median values between groups. *SSc: Systemic Sclerosis*

Demographic, clinical, and laboratory data of SSc patients are shown in Table 2. The disease duration of SSc patients ranged from 1-to 36 years (mean 13.01±8.7). 43 (76.7%) SSc patients had diffuse the disease, and 13 (23.3%) had limited disease subtype. Although there was Raynaud’s disease in 28 patients, interstitial lung disease in 32 patients, cardiac involvement in two patients, and pulmonary arterial hypertension in two patients, there was no patient with a history of renal crisis as the involvement pattern. Five patients had overlapping rheumatoid arthritis, Sjogren’s, or myositis accompanying SSc. 55.3% (n:31) of the patients had anti-Scl70 antibodies and 37.5% (n:21) had anti-centromere antibodies. Antibody positivity was not found in 7.2% (n:4) of the patients. SSc patients received different treatments according to their existing complications [hydroxychloroquine 50 (89.2%), colchicine 13 (23.2%), corticosteroids seven (12.5%), pentoxylin 10 (17.8%), 5-phosphodiesterase inhibitor eight (14·2%), endothelin receptor antagonist three (5.3%), acetylsalicylic acid 42 (75%), azathioprine 20 (35.7%), cyclophosphamide five (8.9%)].

**Table 2.** Demographic, Clinical, and Laboratory Data of SSc patients

| Parameters | Value |
| --- | --- |
| Disease duration, mean ± SD [years] | 13.01±8.7 |
| Disease subtype diffuse/limited, n (%) | 43/13 (76.7/23.3) |
| Raynaud's disease, n (%) | 28 (50) |
| Interstitial lung disease, n (%) | 32 (57.1) |
| Cardiac involvement, n (%) | 2 (3.5) |
| Pulmonary arterial hypertension, n (%) | 2 (3.5) |
| Renal crisis, n (%) | 0 (0) |
| Concomitant disease (RA, Sjogren's, myositis), n (%) | 5 (8.9) |
| Anti-Scl70 antibody positivity, n (%) | 31 (55.3) |
| ACA positivity | 21 (37.5) |
| Antibody negative | 4 (7.2) |
| Modified Rodnan Score, mean ± SD | 6.58±2.20 |
| **Medical therapy, n (%)** |  |
| Hydroxychloroquine, n (%) | 50 (89.2) |
| Colchicine, n (%) | 13 (23.2) |
| Corticosteroids, n (%) | 7 (12.5) |
| Calcium channel blocker, n (%) | 38 (67.8) |
| Pentoxifylin, n (%) | 10 (17.8) |
| 5-phosphodiesterase inhibitor, n (%) | 8 (14.2) |
| Endothelin receptor antagonist, n (%) | 3 (5.3) |
| Acetylsalicylic acid, n (%) | 42 (75) |
| Azathioprine, n (%) | 20 (35.7) |
| Cyclophosphamide, n (%) | 5 (8.9) |

SSc: Systemic Sclerosis, ACA: Anti-centromere Antibody, RA: Rheumatoid Arthritis

Analysis of multiple variances among organ involvement patterns (Raynaud’s disease, interstitial lung disease, cardiac involvement, pulmonary arterial hypertension, renal crisis) among SSc patients, no significant difference was observed between organ involvement patterns and Netrin-1 (p<0.05). In the analysis of multiple variances performed according to the presence of antibodies (anti-Scl70, ACA, antibody-negative) in SSc patients, no significant correlation was found between the presence of antibodies and Netrin-1 levels (p<0.05). There was no significant difference in the Netrin-1 between those with Raynaud’s disease (325.88±208.30) and those without (337.31±209.48) in SSc (p>0.05). There was no significant difference in the Netrin-1 between those with interstitial lung disease (348.28±192.55) and those without (309.36±227.28) (p>0.05). Evaluation in terms of pulmonary arterial hypertension (n:2), cardiac involvement (n:2), and renal crisis (n:0) was not performed because the number of patients was insufficient.

Table 3 shows the ROC analysis results for Netrin-1 in SSc. When the cut-off value of 354.24 was taken for Netrin-1 in the SSc, it was determined that the sensitivity of the test was 32.8%, and the specificity was 98.3%, giving a sensitive confidence interval (AUC [95% Cl]: 0.746-0.895, p<0.0001). Figure 2 shows the ROC analysis graph of Netrin-1 levels.

**Table 3.** Specificity, Sensitivity, and the Cut-off Levels of Netrin-1 in SSc

|  | Cut-off | AUC(%95 CI) | Sensitivity (%) | Specificity (%) | LR | p-value |
| --- | --- | --- | --- | --- | --- | --- |
| Netrin-1 | 354.24 | 0.821 (0.746-0.895) | 32.8 | 98.3 | 19 | <0.0001 |

*AUC: Area under the ROC Curve, LR: Likelihood ratio, SSc: Systemic Sclerosis*

**Fig. 2.**
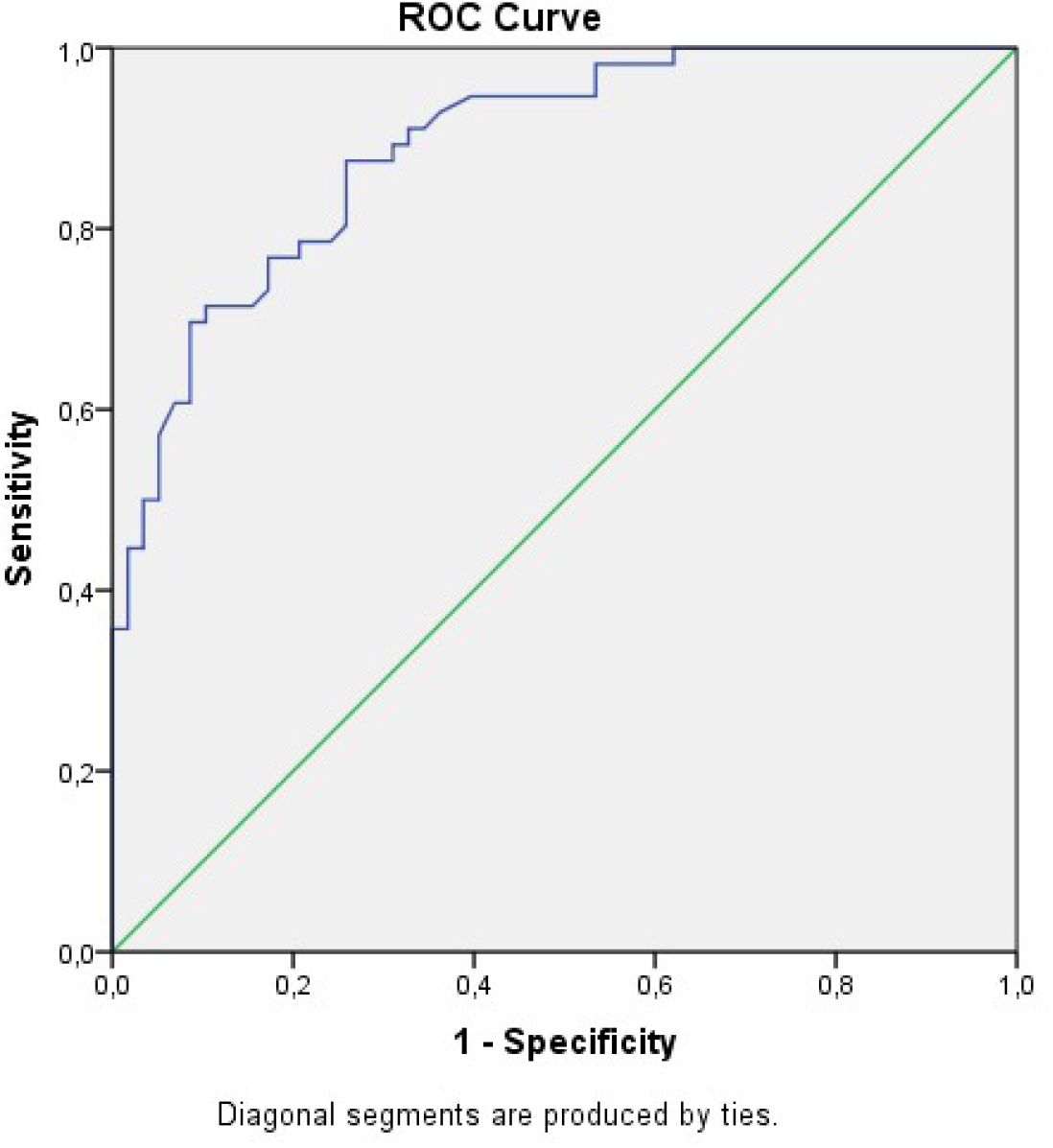
ROC curves of levels of Netrin-1 in Systemic Sclerosis.

Table 4 shows the correlation between study parameters, patient age, and disease duration. A statistically significant positive correlation was found between ESR and patient age (r=0.431, p < 0·01) and disease duration (r=0.847, p<0.01). A negative correlation was found between mRSS and patient age (r=-0.293, p<0.05). No correlation was observed between Netrin-1 and mRSS, CRP, ESR, patient age, and disease duration (p>0.05).

**Table 4.** Correlation Between Study Parameters

| Parameter (r) | Netrin-1 | CRP | ESR | mRSS | Patient age | Disease Duration |
| --- | --- | --- | --- | --- | --- | --- |
| Netrin-1 | - | 0.093 | 0.097 | 0.089 | 0.075 | 0.030 |
| CRP | 0.093 | - | 0.124 | 0.073 | 0.013 | 0.145 |
| ESR | 0.097 | 0.124 | - | -0.040 | 0.431** | 0.847** |
| mRSS | -0.069 | 0.073 | -0.040 | - | -0.293* | -0.55 |
| Patient age | 0.075 | 0.013 | 0.431** | -0.293* | - | 0.535 |
| Disease Duration | 0.030 | 0.145 | 0.847** | -0.55 | 0.535 | - |

*r*: Pearson correlation coefficient, \* $p < 0.05$ , \*\* $p < 0.01$ . CRP: C-reactive protein, ESR: Erythrocyte Sedimentation Rate, mRSS: Modified Rodnan Skin Score

## Discussion

SSc is a progressive disease that pathophysiologically starts with microvascular damage and then develops widespread fibrosis due to increased autoimmune response and inflammation [18]. Dysregulations in vasodilators (nitric oxide [NO], prostacyclins), cell adhesion molecules (e.g. selectins, integrins), and vasoconstrictors are thought to be responsible for the microvascular abnormalities and endothelial dysfunction seen in SSc [19]. In addition, it has been reported that decreased release of vasodilator neuropeptides (serotonin and calcitonin gene-related peptides, etc.) from sensory nerve endings and increased vasoconstrictive responses of vascular smooth muscle α2c adrenoceptors to stimuli contribute to the vasculopathy seen in SSc [20-22].

Limited studies in the literature evaluate the relationship of Netrin-1 with SSc. Two publications evaluate the association with Netrin-1 in bleomycin-induced lung fibrosis in mice and SSc lung cell culture in humans. Ruijuan G. et al. showed that mouse lung fibrosis due to bleomycin is stimulated by macrophage-derived Netrin-1 concerning the adrenergic nerve [16]. In a study conducted in human SSc lung cell culture, it was shown that Netrin-1 expression increased in SSc lung cells. This study also found that bleomycin-induced lung fibrosis and fibrocyte accumulation did not occur in the absence of Netrin-1 in mice [17]. However, no study evaluated Netrin-1 plasma levels in the SSc patient population. This study investigated the relationship between SSc disease and Netrin-1 plasma levels. In the results we obtained, we found that the median value of Netrin-1 was significantly higher in SSc than in healthy controls (p<0.0001). In our study, while Netrin-1 was not found to be associated with SSc organ involvement and skin firmness (p>0.05), we found that it has a high specificity (98.3%) diagnostic value in SSc (AUC[95% Cl]: 0.746-0.895, p<0.0001).

It has been reported in the recent literature that VEGF-A, a proangiogenic factor, plays a role in the pathophysiology of SSc. However, although it has been shown that there is an increase in VEGF-A in the serum and skin tissue samples of patients with SSc, it is also known that there is angiogenesis deficiency in SSc [23]. Studies have determined that VEGF165b antiangiogenic isoform and VEGF165 proangiogenic isoform are found due to alternative splicing in VEGF-A pre-mRNA [24,25]. It has been reported that VEGF165b is stored at high levels in platelets in SSc and may cause high serum VEGF165b levels by activating the platelets by contacting the damaged SSc endothelium [26]. In a study examining the dermis of the SSc, it was reported that VEGF165b antiangiogenic isoform was overexpressed in fibroblasts, endothelial, and perivascular mononuclear inflammatory cells. The elevation in VEGF165b levels correlated with vascular loss in nail capillaroscopy [27]. In the results of the research; it has been determined that Netrin-1 increases both VEGF expression and VEGF receptor response. In animal studies in which experimental limb ischemia was created, it was shown that VEGF expression was significantly higher in subjects who received Netrin-1 treatment compared to those who did not (p<0.01) [13,14]. Similarly, in a study by Park et al., it was found that Netrin-1 is a potent vascular mitogen. It increases VEGF, VEGF receptor level, and VEGF receptor responses [15]. However, although it has been reported that Netrin-1 increases VEGF in these studies, it has not been evaluated which VEGF isoforms it increases. Further research studies evaluating the association of Netrin-1 with VEGF in SSc are required. Detection of the role of VEGF165b increase due to Netrin-1 in the pathophysiology of angiogenesis failure in Ssc seems interesting.

Macrophages formed by monocyte differentiation can have two cell phenotypes classified according to the markers on the cell surface: Classically activated (M1) and alternatively activated (M2) form [28]. M1 macrophages are generally accepted as effector phagocytic cells that can produce microbicidal or tumoricidal effects by secreting proinflammatory cytokines such as IL-1, IL-6, and TNF-alpha. M2 polarized macrophages, which have anti-inflammatory properties, are generally involved in the synthesis of cytokines such as IL-13, IL-10, and IL-4 [29]. As inducers of tissue fibrosis in SSc, M2 macrophages act at the apex of the profibrotic late immune response or during wound healing of tissues. It has been reported that M2 macrophages increase extracellular matrix (ECM) protein synthesis and profibrotic cytokine synthesis, partially suppress M1-induced inflammation, and strengthen the anti-inflammatory response by inducing Th2 effector cell activity [29]. Similarly, in another study on gastric malignancy, M2 macrophages were found to cause ECM increase [30]. Interestingly, the results from studies show that M1 macrophage marker expression is suppressed due to Netrin-1, while M2 macrophage marker expression is increased [31-33]. These results suggest that Netrin-1 may be a possible candidate mediating the known effects of M2 macrophages on ECM increase in SSc.

The most important limitation of our study is that it is a cross-sectional study, and only a small number of patients were included. However, this study showed that the increase in Netrin-1 plasma levels is closely related to SSc disease. Considering the known roles of VEGF and M2 macrophages in the development of SSc and their close relationship with Netrin-1, further studies in this area seem to create a fertile field in understanding the complex pathophysiology of SSc and offering new therapeutic options. The effects of anti-Netrin-1 targeted therapies on the progression of fibrosis in SSc via M2 macrophages, and VEGFR need to be investigated further.

## Conclusion

The median value of Netrin-1 was found to be significantly higher in SSc (268.8 [82.75-1006.64]) than in controls (108.63 [21.02-351.49]) (AUC[95% CI]: 0.746-0.895), p<0.0001). Our study is the first clinical study in which Netrin-1 elevation was demonstrated in SSc patients.

## Data Availability

All data produced in the present study are available upon reasonable request to the authors

https://www.ncbi.nlm.nih.gov

## Contributors

YM, AK, EFO, and KG collected laboratory data; YM, AK, KG, AS and AA collected and analyzed clinical data; YM, AK designed the study; Data analysis and interpretation: all authors; AK wrote the article; Article draft and revision: YM, AK, AA

## Acknowledgments

This study was funded by the Turkish Rheumatology Association (grant: 01.02.2021). We thank the patients who participated in the study that this study could not be done without their support. This study was conducted in compliance with the Declaration of Helsinki and the principles of Good Clinical Practice and standard operating procedures that ensure compliance with all applicable legal requirements.

## Ethics approval

The ethics committee approval of the research protocol was made by the Ankara City Hospital Ethics Committee. Protocol number: E1-20-406 Informed consent was obtained from the patients to participate in the study.

## Patient consent for publication

Not required

## Data sharing statement

All data requests should be submitted to the corresponding author (AK) for consideration as agreed in our publication plan. The article may be used, downloaded, and printed for any lawful, non-commercial purpose (including text and data mining), with all copyright notices and trademarks reserved.

